# Non-alcoholic fatty liver disease in the South Asian Region: A systematic review and meta-analysis

**DOI:** 10.1101/2022.05.30.22275764

**Authors:** Madunil Anuk Niriella, Dileepa Senajith Ediriweera, Madhuri Yasodha Withanage, Selani Darshika, Shamila Thivanshi De Silva, Hithanadura Janaka de Silva

## Abstract

**Objectives:** We estimated the prevalence and effect sizes of associations for NAFLD, the commonest chronic liver disease worldwide, among South-Asian adults.

**Design:** We searched the PubMed database, using search terms “Prevalence of NAFLD (Non-Alcoholic Fatty Liver Disease)” AND “South Asia” AND South Asian countries (“Afghanistan”, “Bangladesh”, “Bhutan”, “India”, “Maldives”, “Nepal”, “Pakistan” and “Sri Lanka”). We included descriptive, epidemiological studies with satisfactory methodology, reporting the prevalence of NAFLD with a valid diagnostic method (ultrasound/CT imaging, biochemistry, histology). The quality of the studies was assessed using Joanna Briggs Institute Critical Appraisal Checklist for Prevalence Studies. Two authors screened and extracted data independently. A random-effects meta-analysis of prevalence and effect sizes of associations of NAFLD was performed. Gender, urban/rural setting, general population and individuals with metabolic diseases (MetD) stratified the analysis.

**Results:** Thirty-two articles were included in the systematic review, and 21 publications were included in the meta-analysis after quality assurance. The pooled overall prevalence of NAFLD in the general population was 25.2% [95%CI 20.3-30.5%] with high heterogeneity (k=9; Q=251.6, DF=8, P<0.0001, I^2^=96.8%). The prevalence was similar among men and women (Q=0.10, DF=1, P=0.746). The NAFLD prevalence in the rural communities were 26.0% (95%CI: 18.2–34.5%) and it was 26.6% (95%CI: 20.5-33.1%) in urban communities without significant differences in the prevalence (Q=0.01, DF=1, P= 0.916). The pooled overall prevalence of NAFLD in patients with MetD was 55.1% [95%CI 47.4-62.8%] with high heterogeneity (k=8; Q=53.8, DF=7, P<0.0001, I^2^=85.2%).

The pooled overall prevalence of NAFLD in the non-obese population was 11.7% [95%CI 7.0-17.3%] (k=6; Q=170.1, DF=5, P<0.0001; I2=97.1%). The pooled prevalence of non-obese NAFLD in the NAFLD population was 43.4% [95%CI 28.1-59.4%] [k=6; Q=181.1; P<0.0001; I2=97.2%]. Meta-analysis of binary outcomes showed presence of NAFLD in South Asian population was associated with diabetes mellitus [RR-2.03 (1.56-2.63)], hypertension [RR-1.37 (1.03-1.84)], dyslipidaemia [RR-1.68 (1.51-1.88)], general obesity [RR-2.56 (1.86-3.51)], central obesity [RR-2.51 (1.69-3.72)] and metabolic syndrome [RR-2.86 (1.79-4.57)]. Gender was not associated with NAFLD.

**Conclusions:** The overall prevalence of NAFLD among adults in South Asia is high, especially those with metabolic abnormalities, and a considerable proportion are non-obese. In the South Asian population, NAFLD was associated with MetD.

**Strengths and limitations:**

- There has been no meta-analysis of epidemiological data on NAFLD from the South Asian region.
- Therefore, we estimated the overall prevalence and effect sizes of risk factors for NAFLD among South-Asian adults.
- We carried out an extensive quality assessment of the studies and included only studies with satisfactory methodological quality in the final analysis to ensure the validity of the results.
- In the present study, the prevalence of NAFLD among adults in the South Asian region seems compatible with the global average, and the prevalence was especially in individuals with metabolic abnormalities.
- This study was limited to available data among adults, excluding the paediatric and adolescent population.

## Introduction

Non-alcoholic fatty liver disease is the commonest chronic liver disease worldwide [1]. It is characterised by hepatic steatosis detected by either imaging or histology without secondary causes. It spans a spectrum of the disease from non-alcoholic fatty liver (NAFL) to non-alcoholic steatohepatitis (NASH) and ultimately to cirrhosis and its complications [2]. Most patients have simple fatty liver with no or mild non-specific inflammation, without liver fibrosis. Conversely, NASH, the more active form of the disease, has varying degrees of liver fibrosis, resulting in cirrhosis and its complications and comorbid cardiovascular disease [3].

Recent studies report the global pooled prevalence of NAFLD at 25.2%, with wide geographical variation worldwide.[1] The highest prevalence rates of up to 30%, primarily based on abdominal ultrasound, are from the Middle East and South American countries [4]. There is an association between NAFLD prevalence and overweight and diabetic status [5]. Whilst a large part of the worldwide increase in NAFLD is driven by obesity,[4] patterns of increased prevalence do not always associate with areas of higher caloric consumption, suggesting that other factors may be contributing to the progression of NAFLD[6, 7].

The prevalence of NAFLD in South Asian countries is high [8, 9]. This is presumably due to several factors, which include socioeconomic growth, urbanisation, westernised diet, increasingly sedentary lifestyle and poor health awareness [9]. Almost 20% of the world’s population resides in South Asia, which renders it the most densely populated region with a large number of patients with NAFLD and metabolic syndrome [9].

There have been two previous systematic reviews on NAFLD from the South Asian region.[8, 9] These studies evaluated NAFLD’s prevalence, associations, and phenotype in South Asian countries, including Bangladesh, India, Nepal, Pakistan, and Sri Lanka. However, there has been no meta-analysis of epidemiological data on NAFLD from this region.

In the present paper, studies on the prevalence of NAFLD and its associations in South Asian countries up to October 2021 were searched and included in a systematic review and subsequent meta-analysis. The aim was to estimate the overall prevalence and effect sizes of the associated risk factors for NAFLD among adults in the South Asian region.

## Material and methods

### Design

We followed the preferred reporting items for systematic reviews and meta-analysis (PRISMA) guidelines for the conduct of this study (Fig. 1).

**Fig. 1.**
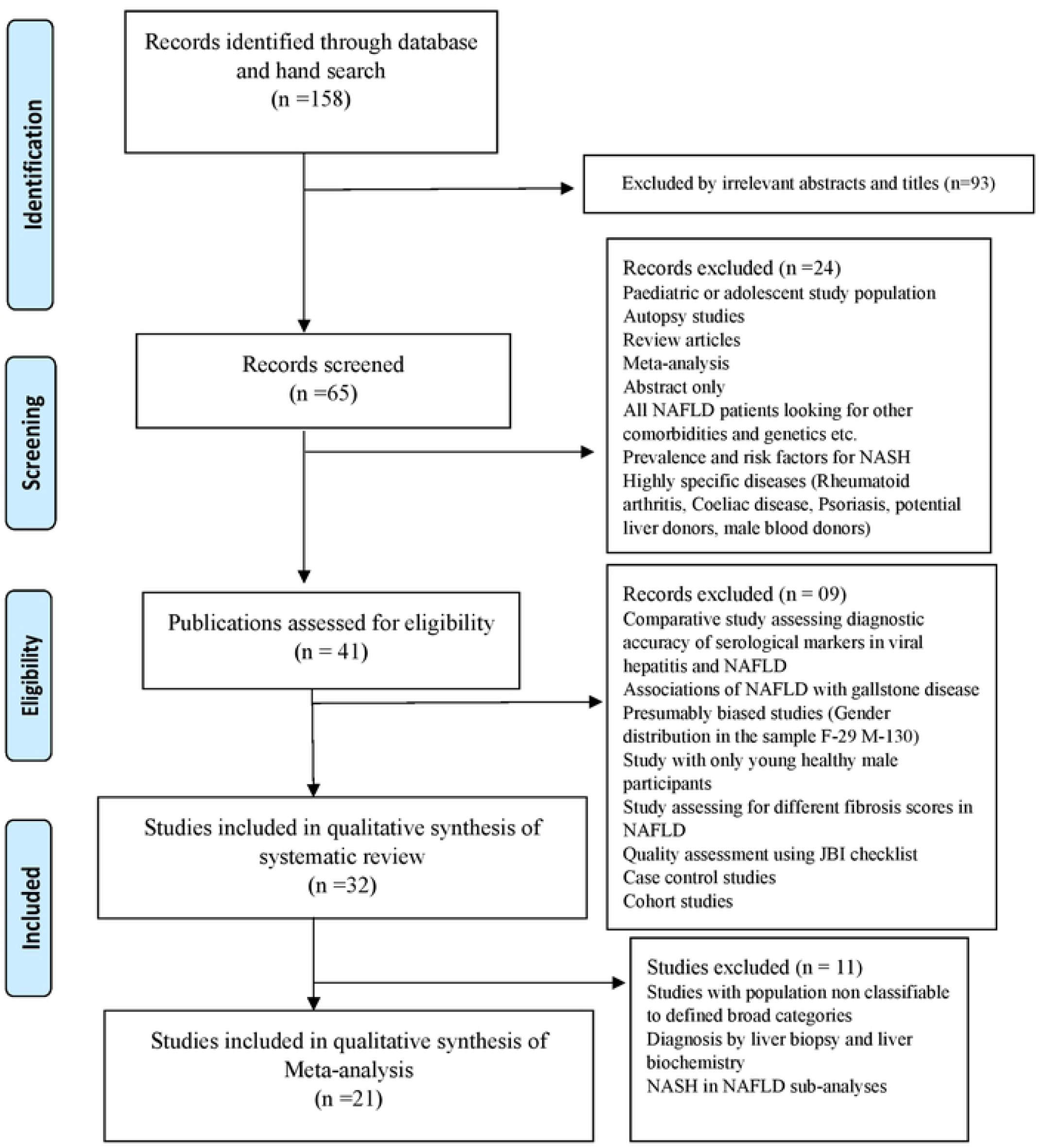
PRISMA flowchart for selection of studies

### Literature survey

A comprehensive survey on PubMed was carried out for articles published up to October 2021 with information on the prevalence, associations and risk factors for NAFLD in South Asia. The search terms included “Prevalence of NAFLD (Non-Alcoholic Fatty Liver Disease)” AND “South Asia” AND individual South Asian countries (“Afghanistan”, “Bangladesh”, “Bhutan”, “India”, “Maldives”, “Nepal”, “Pakistan” and “Sri Lanka”).

### Inclusion and exclusion criteria

The following inclusion criteria were used to select eligible articles published before October 2021: (1) descriptive, epidemiological studies reporting NAFLD, (2) studies conducted in the South Asian region (Afghanistan, Bangladesh, Bhutan, India, Maldives, Nepal, Pakistan and Sri Lanka), (3) studies including adults 18 years of age and over, (4) studies that confirmed NAFLD with a valid diagnostic method (ultrasound or other imaging, liver biochemistry, histology) and (5) studies published in the English language. In addition, we also included studies reporting NAFLD prevalence among participants with metabolic conditions, such as morbid obesity, diabetes mellitus, metabolic syndrome and polycystic ovarian disease. The exclusion criteria were: (1) articles in a non-English language, (2) review articles and guidelines and (3) clinical trials.

### Selection of studies

Two authors (MAN, MYW) screened the title and abstract of all the studies identified to select potentially eligible studies for the systematic review and meta-analysis. Next, bibliographies of the chosen articles, specifically the available relevant review articles, were cross-checked and searched by hand to possibly identify additional studies. Finally, full texts of eligible articles were retrieved and studied in depth. The final decision on the inclusion of an article was in line with the agreed criteria, and disagreements and disparities during the process were resolved by a third author (DSE).

Articles selected for the systematic review were further evaluated for the synthesis of meta-analysis in two main steps: pooling the prevalence of NAFLD and pooling the effect sizes of NAFLD associations (Fig. 1). In the first step (pooling the prevalence of NAFLD), descriptive studies and case-control studies reporting the NAFLD prevalence were selected. After that, the studies were divided into sub-analyses, where possible, by the setting (as “urban” or “rural”) of the general population and by obese and non-obese populations in the studies focusing on obese and non-obese NAFLD. Missing data for one study [10] was extracted from the relevant prevalence study on NAFLD in the same cohort [11]. Exclusion criteria for the meta-analysis were: studies where NAFLD was diagnosed by methods other than ultrasound (liver biochemistry or histology), case-control studies, studies of NASH in NAFLD populations and studies which precluded the classification of participants into required categories or settings. In the second step (pooling the effect sizes of NAFLD associations), available data for associations of NAFLD were extracted from the selected studies.

### Data extraction

Following data were extracted from the studies: author name, year of publication, study designs, sample size, age group of participants (>18 years), study setting, diagnostic criteria used for diagnosing NAFLD, characteristics of the study population and risk category of the participants. Data from selected studies were extracted for the estimation of pooled rates for the prevalence of NAFLD and associations of NAFLD. Two authors performed data extraction independently (MAN, MYW), and any discrepancies were resolved by discussion with a third author (DSE).

Based on the available data, the selected studies were categorised as having a “general population” or “individuals having metabolic diseases”. The “general population” studies had chosen participants from the community with a presumed “average risk” of NAFLD. On the other hand, studies with “individuals having metabolic diseases” had participants with a metabolic derangement (diabetes mellitus/pre-diabetes mellitus, obesity, metabolic syndrome, cardiovascular disease, women with polycystic ovarian disease) with presumed “high risk” for NAFLD. The studies that were identified as “general population” were further divided into “rural” and “urban”.

Factors associated with NAFLD were extracted from the selected articles, after which, frequently reported factors were considered for pooling in the second step of the study. The selected factors were: male gender, general obesity, central obesity, diabetes mellitus, dysglycaemia, hypertension, dyslipidaemia and metabolic syndrome. Since studies reported different forms of glycaemic abnormalities, the term “dysglycaemia” was also used to combine diabetes mellitus, hyperglycaemia and insulin resistance. The cut-off values for dyslipidaemia varied across the selected articles. Therefore, participants who were already categorised as having hypertriglyceridemia/ dyslipidaemia/high triglyceride (TG) levels or low high-density lipoproteins (HDL) on treatment/TG levels above 150 mg/dL or HDL below 40 mg/dL were included in the category of dyslipidaemia. Varying definitions have been used for general obesity. Therefore, participants were directly categorised as obese in the study and participants with a body mass index (BMI) above the Asia-Pacific cut-off value (>25kgm^-2^) were included in the obese category. Similarly, participants directly categorised as having central obesity and participants with “high” waist circumference (WC) or participants with WC >90cm in females and >80cm in males were included in the category of central obesity.

### Quality assessment for individual studies

The quality of the studies was assessed using Joanna Briggs Institute Critical Appraisal Checklist for Prevalence Studies [12]. This tool is comprised of 9 questions that consider the following: sample frame appropriateness, recruitment appropriateness, sample size, descriptions of subjects and setting, coverage of data analysis, ascertainment and measurement of the condition, appropriateness of statistical analysis and the adequacy and management of the response rate. Each question had the option to answer ‘yes’, indicating higher quality; ‘no’, indicating poor quality; ‘unclear’ or ‘not applicable. Two authors (SD, DSE) completed quality assessments for the studies considered eligible for inclusion. Any discrepancies in judgements regarding inclusion were resolved through discussion. Based on overall quality, studies with a number of positive responses (‘yes’) greater than six were included in the systematic review and meta-analysis. If a study had ≥ 3 ‘no’ or’ unclear’ quality categories, then it was excluded from the analysis. The outcomes of the appraisal are presented in a table (Table 1).

**Table 1:**
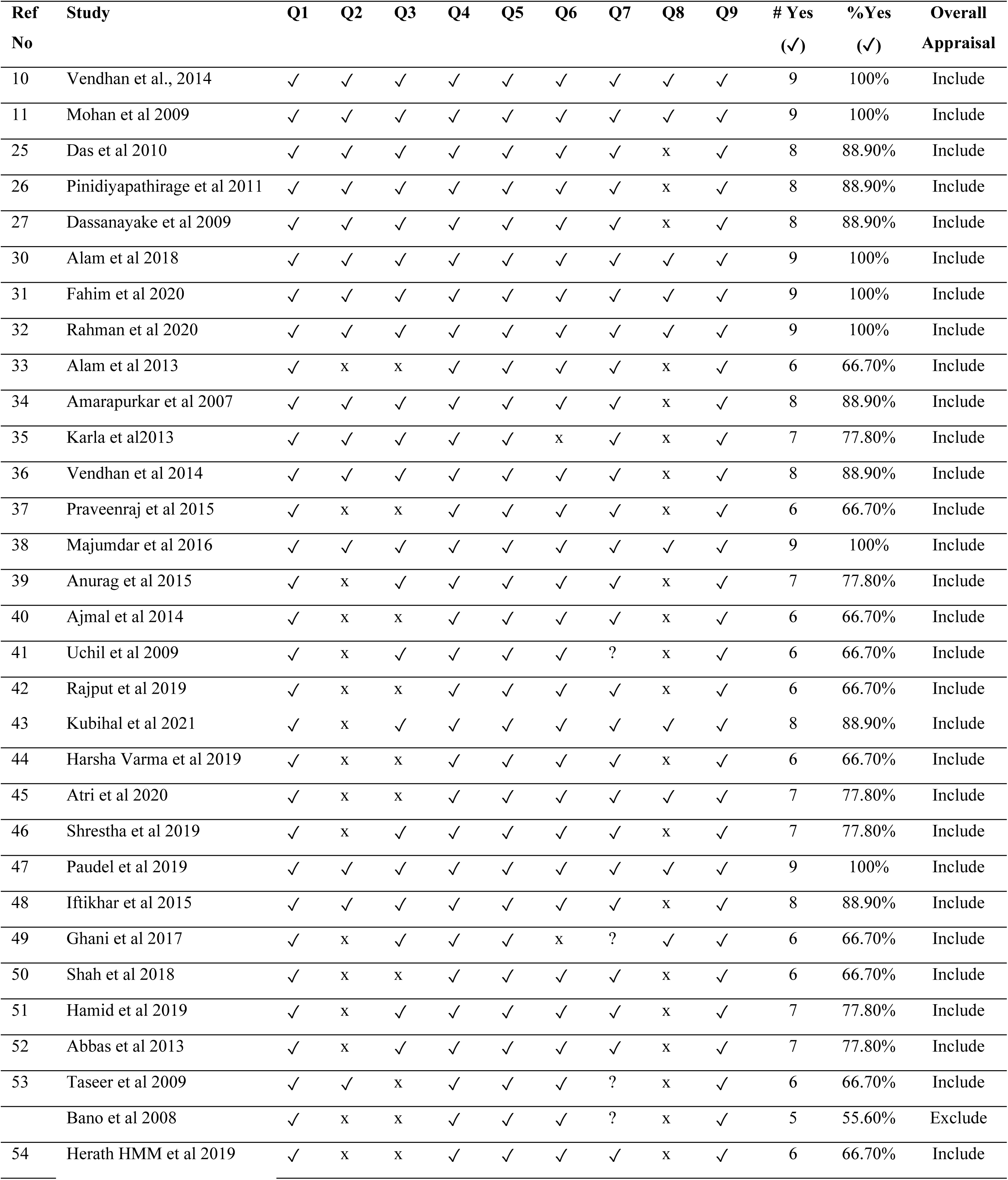
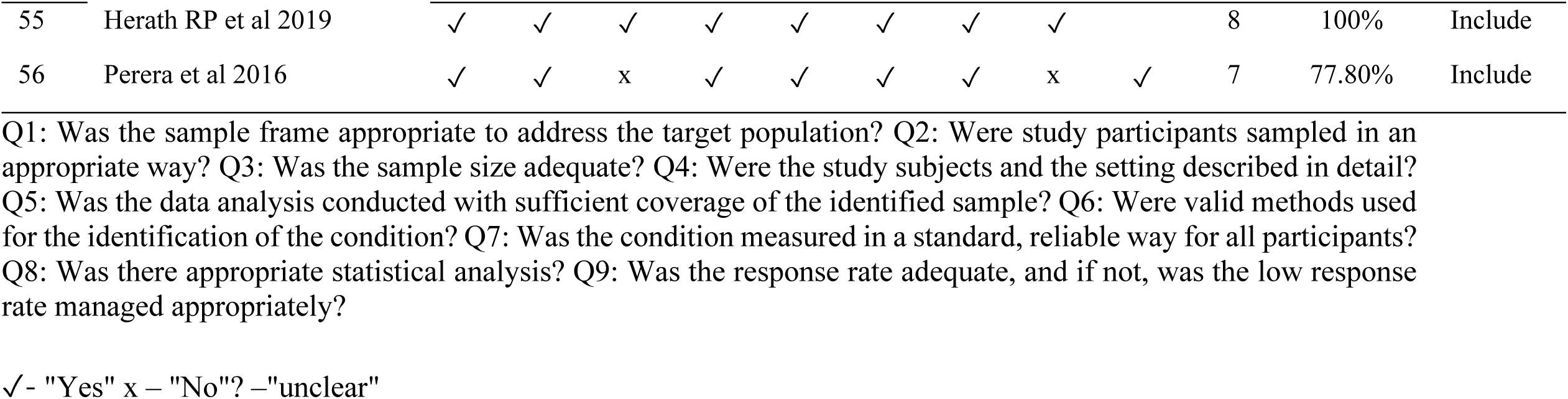
Critical appraisal result using Joanna Briggs Institute (JBI) Critical Appraisal Checklist for Prevalence Studies.

| Ref No | Study | Q1 | Q2 | Q3 | Q4 | Q5 | Q6 | Q7 | Q8 | Q9 | # Yes (✓) | %Yes (✓) | Overall Appraisal |
| --- | --- | --- | --- | --- | --- | --- | --- | --- | --- | --- | --- | --- | --- |
| 10 | Vendhan et al., 2014 | ✓ | ✓ | ✓ | ✓ | ✓ | ✓ | ✓ | ✓ | ✓ | 9 | 100% | Include |
| 11 | Mohan et al 2009 | ✓ | ✓ | ✓ | ✓ | ✓ | ✓ | ✓ | ✓ | ✓ | 9 | 100% | Include |
| 25 | Das et al 2010 | ✓ | ✓ | ✓ | ✓ | ✓ | ✓ | ✓ | x | ✓ | 8 | 88.90% | Include |
| 26 | Pinidiyapathirage et al 2011 | ✓ | ✓ | ✓ | ✓ | ✓ | ✓ | ✓ | x | ✓ | 8 | 88.90% | Include |
| 27 | Dassanayake et al 2009 | ✓ | ✓ | ✓ | ✓ | ✓ | ✓ | ✓ | x | ✓ | 8 | 88.90% | Include |
| 30 | Alam et al 2018 | ✓ | ✓ | ✓ | ✓ | ✓ | ✓ | ✓ | ✓ | ✓ | 9 | 100% | Include |
| 31 | Fahim et al 2020 | ✓ | ✓ | ✓ | ✓ | ✓ | ✓ | ✓ | ✓ | ✓ | 9 | 100% | Include |
| 32 | Rahman et al 2020 | ✓ | ✓ | ✓ | ✓ | ✓ | ✓ | ✓ | ✓ | ✓ | 9 | 100% | Include |
| 33 | Alam et al 2013 | ✓ | x | x | ✓ | ✓ | ✓ | ✓ | x | ✓ | 6 | 66.70% | Include |
| 34 | Amarapurkar et al 2007 | ✓ | ✓ | ✓ | ✓ | ✓ | ✓ | ✓ | x | ✓ | 8 | 88.90% | Include |
| 35 | Karla et al 2013 | ✓ | ✓ | ✓ | ✓ | ✓ | x | ✓ | x | ✓ | 7 | 77.80% | Include |
| 36 | Vendhan et al 2014 | ✓ | ✓ | ✓ | ✓ | ✓ | ✓ | ✓ | x | ✓ | 8 | 88.90% | Include |
| 37 | Praveenraj et al 2015 | ✓ | x | x | ✓ | ✓ | ✓ | ✓ | x | ✓ | 6 | 66.70% | Include |
| 38 | Majumdar et al 2016 | ✓ | ✓ | ✓ | ✓ | ✓ | ✓ | ✓ | ✓ | ✓ | 9 | 100% | Include |
| 39 | Anurag et al 2015 | ✓ | x | ✓ | ✓ | ✓ | ✓ | ✓ | x | ✓ | 7 | 77.80% | Include |
| 40 | Ajmal et al 2014 | ✓ | x | x | ✓ | ✓ | ✓ | ✓ | x | ✓ | 6 | 66.70% | Include |
| 41 | Uchil et al 2009 | ✓ | x | ✓ | ✓ | ✓ | ✓ | ? | x | ✓ | 6 | 66.70% | Include |
| 42 | Rajput et al 2019 | ✓ | x | x | ✓ | ✓ | ✓ | ✓ | x | ✓ | 6 | 66.70% | Include |
| 43 | Kubihal et al 2021 | ✓ | x | ✓ | ✓ | ✓ | ✓ | ✓ | ✓ | ✓ | 8 | 88.90% | Include |
| 44 | Harsha Varma et al 2019 | ✓ | x | x | ✓ | ✓ | ✓ | ✓ | x | ✓ | 6 | 66.70% | Include |
| 45 | Atri et al 2020 | ✓ | x | x | ✓ | ✓ | ✓ | ✓ | ✓ | ✓ | 7 | 77.80% | Include |
| 46 | Shrestha et al 2019 | ✓ | x | ✓ | ✓ | ✓ | ✓ | ✓ | x | ✓ | 7 | 77.80% | Include |
| 47 | Paudel et al 2019 | ✓ | ✓ | ✓ | ✓ | ✓ | ✓ | ✓ | ✓ | ✓ | 9 | 100% | Include |
| 48 | Iftikhar et al 2015 | ✓ | ✓ | ✓ | ✓ | ✓ | ✓ | ✓ | x | ✓ | 8 | 88.90% | Include |
| 49 | Ghani et al 2017 | ✓ | x | ✓ | ✓ | ✓ | x | ? | ✓ | ✓ | 6 | 66.70% | Include |
| 50 | Shah et al 2018 | ✓ | x | x | ✓ | ✓ | ✓ | ✓ | x | ✓ | 6 | 66.70% | Include |
| 51 | Hamid et al 2019 | ✓ | x | ✓ | ✓ | ✓ | ✓ | ✓ | x | ✓ | 7 | 77.80% | Include |
| 52 | Abbas et al 2013 | ✓ | x | ✓ | ✓ | ✓ | ✓ | ✓ | x | ✓ | 7 | 77.80% | Include |
| 53 | Taseer et al 2009 | ✓ | ✓ | x | ✓ | ✓ | ✓ | ? | x | ✓ | 6 | 66.70% | Include |
|  | Bano et al 2008 | ✓ | x | x | ✓ | ✓ | ✓ | ? | x | ✓ | 5 | 55.60% | Exclude |
| 54 | Herath HMM et al 2019 | ✓ | x | x | ✓ | ✓ | ✓ | ✓ | x | ✓ | 6 | 66.70% | Include |
| 55 | Herath RP et al 2019 | ✓ | ✓ | ✓ | ✓ | ✓ | ✓ | ✓ | ✓ |  | 8 | 100% | Include |
| 56 | Perera et al 2016 | ✓ | ✓ | x | ✓ | ✓ | ✓ | ✓ | x | ✓ | 7 | 77.80% | Include |

Q1: Was the sample frame appropriate to address the target population? Q2: Were study participants sampled in an appropriate way? Q3: Was the sample size adequate? Q4: Were the study subjects and the setting described in detail? Q5: Was the data analysis conducted with sufficient coverage of the identified sample? Q6: Were valid methods used for the identification of the condition? Q7: Was the condition measured in a standard, reliable way for all participants? Q8: Was there appropriate statistical analysis? Q9: Was the response rate adequate, and if not, was the low response rate managed appropriately?
✓ - "Yes" x – "No"? – "unclear"

### Statistical analysis and visualisation tools

In South Asia, the prevalence rates of NAFLD, diagnosed by ultrasound, ranged from 8.7% to 73.6%. The data showed differences in the study population (i.e. general and people with metabolic diseases), gender (male and female) and study settings (i.e. urban and rural). Therefore, the overall NAFLD prevalence (pooled estimates) across the studies was determined by performing a random-effects meta-analysis of proportions using the Der Simoniane Laird model. The number of NAFLD patients amongst the total number sampled in each study was considered for the analysis, and inverse variance weighting was used to pool the studies.

Cochran Q test was used to assess the heterogeneity between the studies. Outlier and influential case diagnostics were performed using the externally standardized residuals, DFFITS values, Cook’s distances, covariance ratios, leave-one-out estimates of the amount of heterogeneity, leave-one-out heterogeneity test statistics, hat values and weights[13,14]. Outliers and influential cases were identified and removed before obtaining pooled estimates.

Further, prevalence rates were evaluated for the differences in gender and study setting (i.e. rural versus urban) within general and urban populations. Subsequently, the reported risk factors (i.e., gender, general obesity, central obesity, diabetes mellitus, dysglycaemia, hypertension, dyslipidaemia, and metabolic syndrome) were evaluated to obtain pooled estimates for risk ratios (RR) using random-effects models. This was done by conducting a meta-analysis of binary outcomes where the number of NAFLD cases amongst people with and without the given risk factor was evaluated. Forest plots were developed to summarise the results of the meta-analysis, and the publication bias was assessed and visualised using a funnel plot. A P value of 0.05 was considered significant. Analysis was done using R programming language 3.6.3. A meta-analysis of proportions was carried out using “meta” library. Outliers and influential cases were identified using “dmetar” library. Risk of bias plot and traffic light plots were obtained from “robvis” library.

## Results

### Selection of articles

A total of 158 articles were identified during the initial literature search. We included descriptive, epidemiological studies with satisfactory methodological quality, reporting the prevalence of NAFLD with a valid diagnostic method.

Only 32 publications (four from Bangladesh, 15 from India, two from Nepal, six from Pakistan and five from Sri Lanka) were included in the systematic review after the quality assurance process (Supplementary table 1). Studies included in the systematic review used any valid diagnostic method (ultrasound/CT imaging, liver biochemistry or histology) to determine the presence of NAFLD. The characteristics of the included studies are summarised in Table 2 and supplementary table 1.

**Table 2:** NAFLD in South Asia (up to October 2021)

| Country | Reference | Type | Population category | Mode of diagnosis |
| --- | --- | --- | --- | --- |
| 1) Bangladesh | Alam et al., 2018 [30] | Cross sectional | Urban and rural community | Ultrasound |
| 2) Bangladesh | Fahim et al., 2020 [31] | Cross sectional | Urban and rural community | Ultrasound |
| 3) Bangladesh | Rahman et al., 2020 [32] | Cross sectional | Rural community | Ultrasound |
| 4) Bangladesh | Alam et al., 2013 [33] | Cross sectional | Hospital | Histopathology |
| 5) India | Amarapurkar et al., 2007 [34] | Cross sectional | Urban community | Ultrasound |
| 6) India | Das et al., 2010 [25] | Prospective cohort study | Rural community | Ultrasound & CT |
| 7) India | Kalra et al., 2013 [35] | Cross sectional | Hospital | Biochemical |
| 8) India | Vendhan et al., 2014 [36] | Retrospective cohort study | Hospital | Ultrasound |
| 9) India | Praveenraj et al., 2015 [37] | Retrospective cohort study | Hospital | Biopsy |
| 10) India | Majumdar et al., 2016 [38] | Cross sectional | Rural community | Ultrasound |
| 11) India | Anurag et al., 2015 [39] | Cross sectional | Hospital | Ultrasound |
| 12) India | Ajmal et al., 2014 [40] | Cross sectional | Hospital | Ultrasound |
| 13) India | Mohan et al., 2009 [11] | Cross sectional | Urban community | Ultrasound |
| 14) India | Vendhan., et al., 2014 [10] | Cross sectional | Urban community | Ultrasound |
| 15) India | Uchil et al, 2009 [41] | Cross sectional | Hospital | Ultrasound |
| 16) India | Rajput & Ahlawat, 2019 [42] | Cross sectional | Hospital | Ultrasound |
| 17) India | Kubihal et al., 2021 [43] | Cross sectional | Hospital | Ultrasound |
| 18) India | Harsha Varma et al., 2019 [44] | Cross sectional | Hospital | Ultrasound |
| 19) India | Atri et al., 2020 [45] | Cross sectional | Hospital | Ultrasound |
| 20) Nepal | Shrestha et al., 2019 [46] | Cross sectional | Hospital | Ultrasound |
| 21) Nepal | Paudel et al., 2019 [47] | Cross sectional | Urban community | Ultrasound |
| 22) Pakistan | Iftikhar et al., 2015 [48] | Cross sectional | Hospital | Ultrasound |
| 23) Pakistan | Ghani et al., 2017 [49] | Cross sectional | Hospital | Biochemical |
| 24) Pakistan | Shah et al., 2018 [50] | Prospective cohort study | Hospital | Ultrasound & Biopsy |
| 25) Pakistan | Hamid et al., 2019 [51] | Cross sectional | Hospital | Ultrasound |
| 26) Pakistan | Abbas et al., 2013 [52] | Cross sectional | Urban community | Ultrasound |
| 27) Pakistan | Taseer et al., 2009 [53] | Cross sectional | Hospital | Ultrasound |
| 28) Sri Lanka | Pinidiyapathirage et al., 2011 [26] | Cross sectional | Rural community | Ultrasound |
| 29) Sri Lanka | H. M. M. Herath et al., 2019 [54] | Prospective cohort study | Hospital | Ultrasound |
| 30) Sri Lanka | R. P. Herath et al., 2019 [55] | Cross sectional | Hospital | Ultrasound |
| 31) Sri Lanka | Dassanayake et al., 2009 [27] | Prospective cohort study | Urban community | Ultrasound |
| 32) Sri Lanka | Perera et al., 2016 [56] | Cross sectional study | Hospital | Ultrasound |

Only 21 publications (3 from Bangladesh, 11 from India, four from Sri Lanka, four from Pakistan and one from Nepal) were included in the subsequent meta-analysis after the quality assurance process. Studies included in the meta-analysis used only ultrasound/CT imaging to determine the presence of NAFLD. The meta-analysis included 15,455 study participants and 4,741 NAFLD cases. Two of the studies had only female participants, while one had only male participants. There were 7,280 males, including 2062 NAFLD cases and 7,209 females, including 2035 NAFLD cases.

### Assessment of methodological quality

The quality assessment of the selected studies for the meta-analysis is presented in Fig. 2 and Fig. 3. The majority of the studies had a low risk of bias in each of the nine domains of the Joanna Briggs Institute Prevalence Critical Appraisal Tool, 2014 [12].

**Fig. 2.**
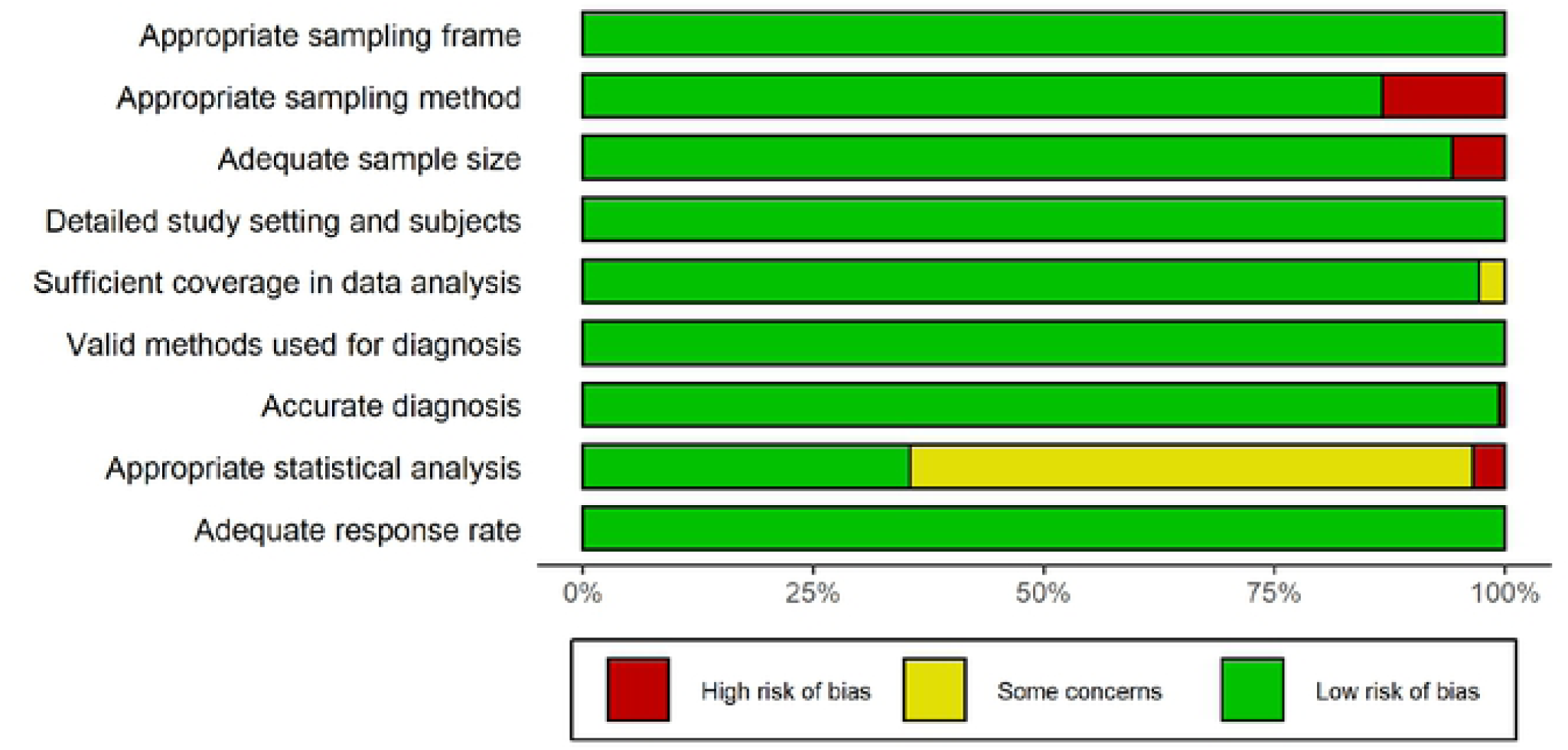
Risk of bias graph: Review of authors’ judgments regarding each risk-of-bias item presented as percentages across included studies

**Fig. 3.**
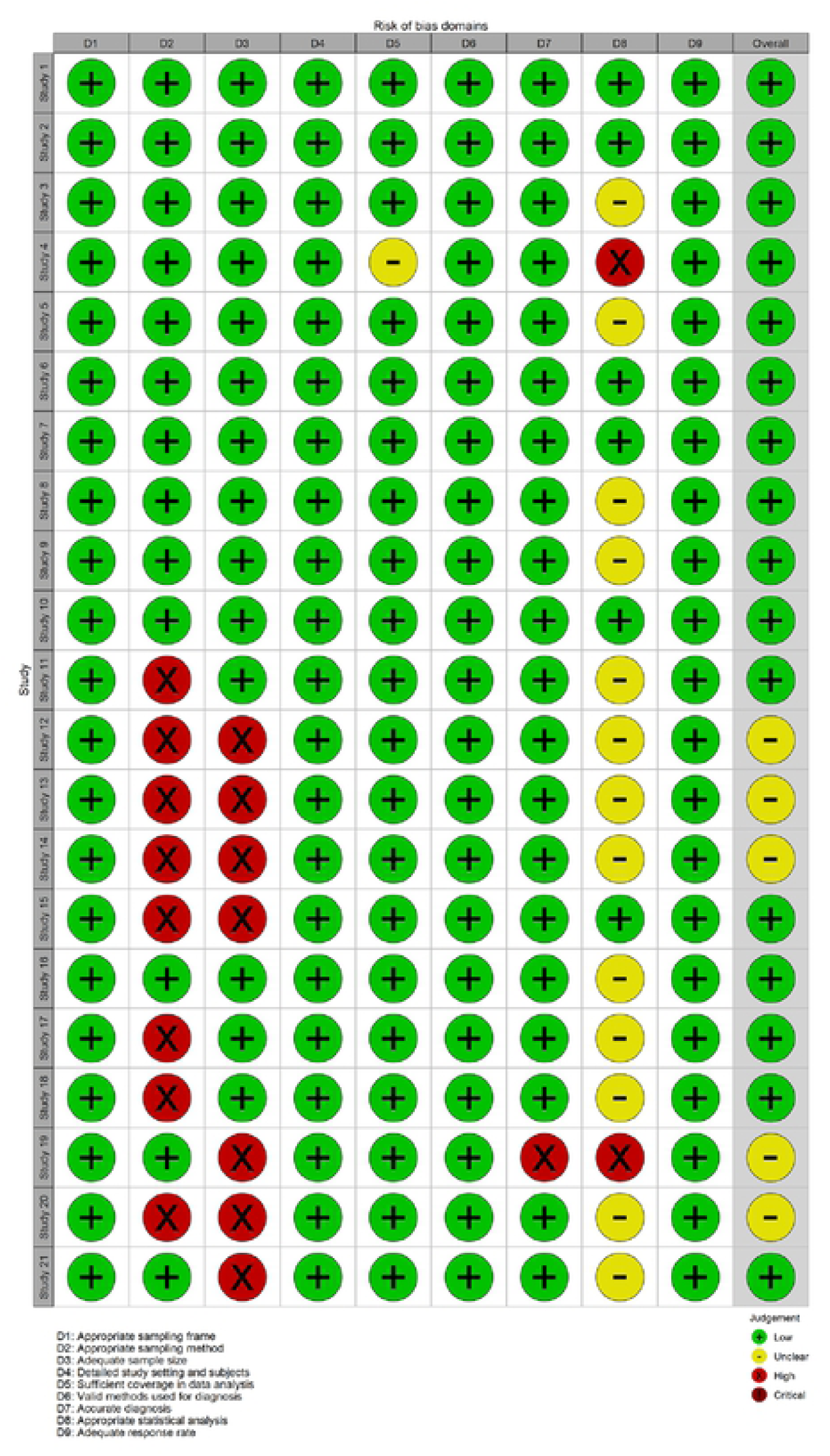
Risk-of-bias summary: The authors’ judgments regarding each risk-of-bias item for each included study

### Heterogeneity of selected studies

Meta-analysis of the prevalence rates showed a high heterogeneity (I^2^=98.8%, Q=1845.36, DF=22, P<0.001). The overall NAFLD prevalence was 40.4% (95%CI: 33.1% – 47.9%) when all the studies were considered together. There were 11 studies representing the “general population” (12,512 participants and 3,374 NAFLD cases) and 12 studies from “individuals with metabolic disease” (2,943 participants and 1,367 NAFLD cases), including three studies with overweight or obese NAFLD populations and one with a morbidly obese population. Subgroup analysis showed significant difference in the prevalence rates between the general population and people with metabolic diseases (Q = 15.8, DF = 1, P < 0.001). Therefore, a separate analysis was conducted for the general population and people with metabolic diseases (Supplementary figure 1).

### Prevalence of NAFLD in the general population

Eleven studies reported NAFLD prevalence rates in the general population from Bangladesh, India, Nepal, Pakistan and Sri Lanka [11,25,26,27,30,32,34,38,39,47,52]. Das et al., 2010 [25] and Paudel et al., 2019 [47] were identified as outliers considering the influence on pooled effect sizes and contribution to heterogeneity in prevalence rates (Supplementary figure 2).

After removing the above two outliers, 10028 individuals were considered in the meta-analysis, including 2822 NAFLD cases. The pooled overall prevalence of NAFLD in the general population was estimated as 25.2% [95% CI 20.3-30.5] (Fig. 4). The prevalence rates showed high heterogeneity (k=9; Q=251.6, DF=8, P<0.0001, I^2^=96.8%).

**Fig. 4.**
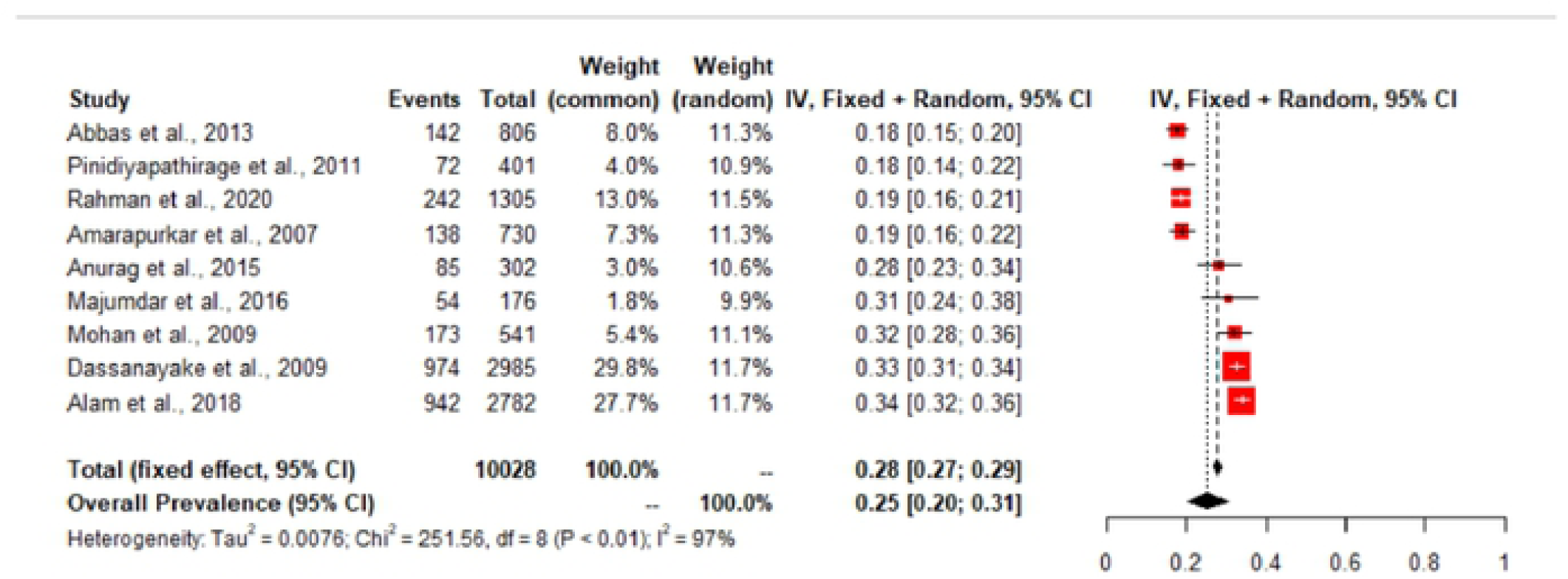
Forest plot of NAFLD prevalence rates in the general population

The general population included 4921 males and 5107 females. The overall NAFLD prevalence among males was 26.0 (95% CI : 21.8% -30.3%) and among females was 24.7% (95% CI : 17.8% - 32.0%). The subgroup analysis did not show gender difference in the NAFLD prevalence rates (Q=0.10, DF = 1, P=0.746) (Supplementary figure 3).

Among eleven studies reporting NAFLD prevalence rates in the general population, five studies provided rural and urban settings data. There were 2848 participants in rural settings and 7180 participants in urban settings. The NAFLD prevalence in the rural communities were 26.0 (95%CI: 18.2 – 34.5%) and it was 26.6 (95%CI: 20.5 - 33.1%) in urban communities without significant differences in the prevalence rates (Q=0.01, DF=1, P= 0.916) (Supplementary figure 4).

### Prevalence of NAFLD in those with metabolic disease

Twelve studies reported NAFLD prevalence rates in patients with metabolic disorders from Bangladesh, India, Pakistan and Sri Lanka [10,32,36,40,42,44,45,48,51,53,54,56]. Vendhan et al., 2014 [36], Rahman et al., 2020 [32], Atri et al., 2020 [45] and Hamid et al., 2019 [51] were identified to have influenced pooled effect sizes and contributed to heterogeneity (Supplementary figure 5).

After removing the above four outliers, 1397 individuals were considered for the analysis, including 754 NAFLD cases. The pooled overall prevalence of NAFLD in patients with metabolic diseases was 55.1% [95% CI 47.4 - 62.8] (Fig. 5). The prevalence rates showed high heterogeneity (k=8; Q=53.8, DF=7, P<0.0001, I^2^=85.2%).

**Fig. 5.**
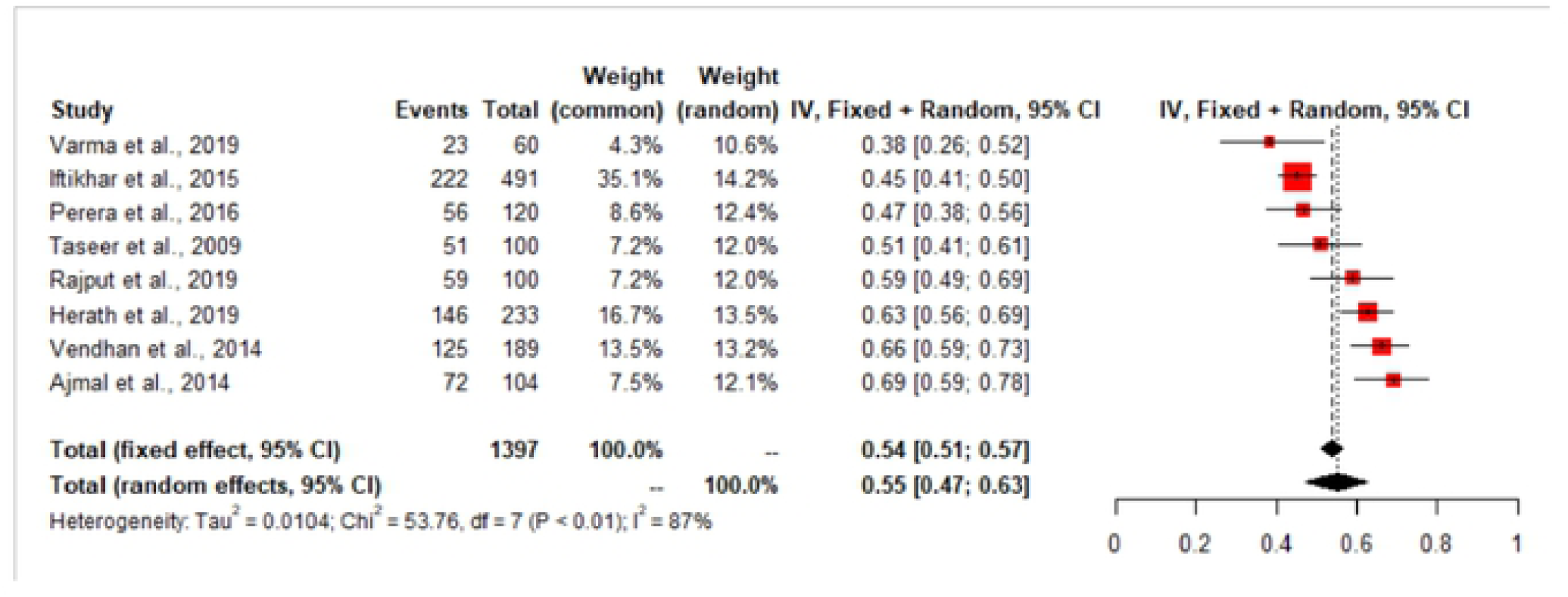
Forest plot of NAFLD prevalence rates in the population with metabolic diseases

From these studies, data were available for 725 males and 279 females. Note that all the studies did not have males and females (Varma et al., 2019, Taseer et al., 2009, Herath et al., 2019, Perera et al., 2016 provided data for females and Iftikhar et al., 2015, Taseer et al., 2009, Herath et al., 2019, Perera et al., 2016 provided data for males). The subgroup analysis showed the overall NAFLD prevalence was 46.8 (95%CI : 37.8 - 55.9%) in males and was 56.3% (95% CI : 43.8 - 68.4%) in females, without significant gender difference in the prevalence rates (Q = 1.44, DF = 1, P=0.229) (Supplementary figure 6).

### Non-obese NAFLD

Six studies provided data on non-obese NAFLD. This included 5612 non-obese participants and 651 non-obese NAFLD cases. The pooled overall estimate for prevalence of NAFLD in the non-obese population was 11.7% [95% CI 7.0-17.3%] (k=6; Q=170.1, DF = 5, P<0.0001; I2 = 97.1%, Figure 3a). These six studies included a total 1767 NAFLD cases. The pooled estimate for the prevalence of non-obese NAFLD in the NAFLD population was 43.4% [95% CI 28.1-59.4%] [k = 6; Q=181.1; P<0.0001; I2= 97.2%] (Fig. 6).

**Fig. 6:**
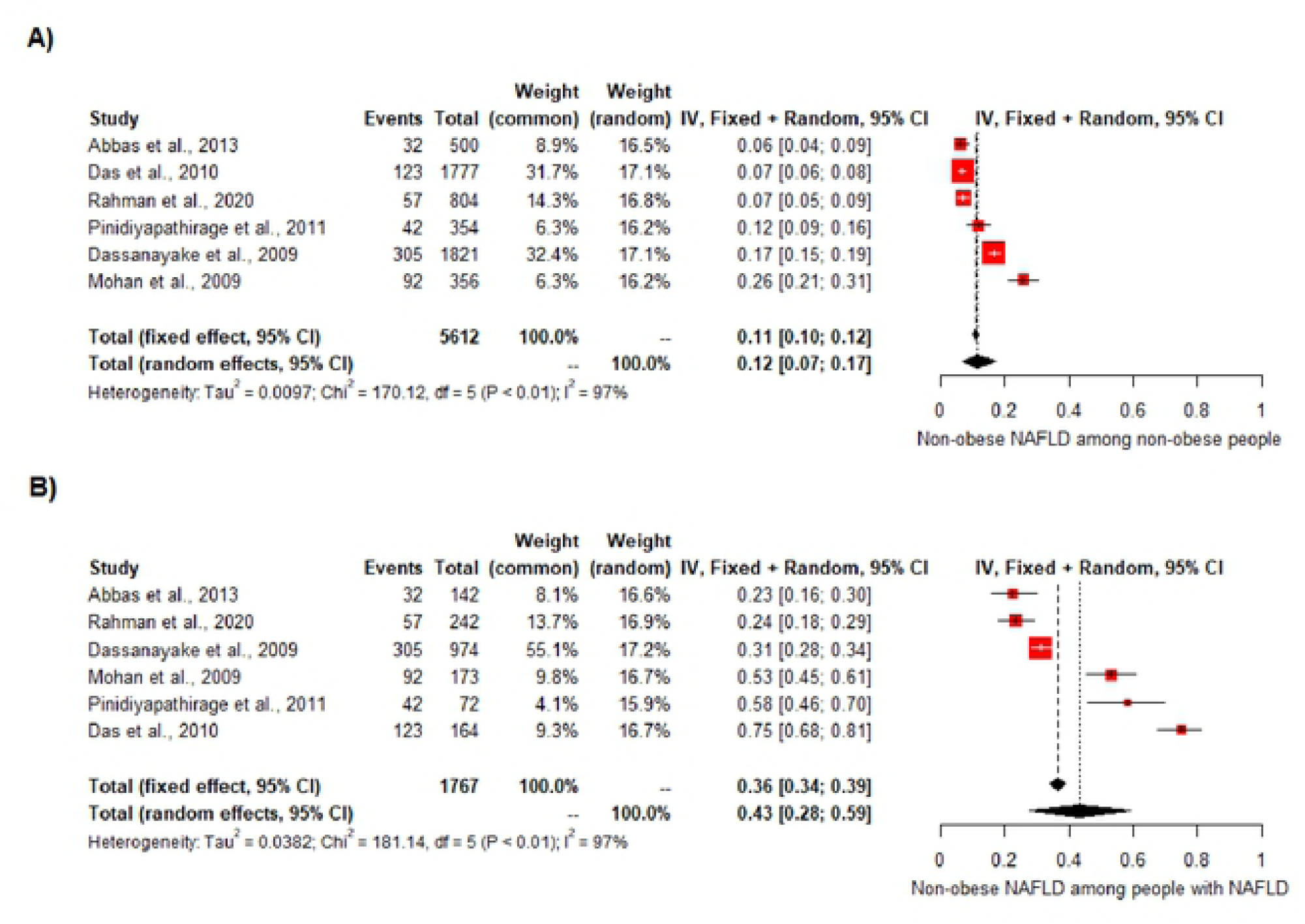
Non-obese NAFLD in A) non-obese population and B) NAFLD population

### Associations for NAFLD

Meta-analysis of binary outcomes showed the presence of NAFLD in the South Asian population was associated with diabetes mellitus, hypertension, dyslipidaemia, general obesity, central obesity and metabolic syndrome. Gender was not associated with NAFLD. The estimated Relative Risk (RR) for each associated factor from random effect models is shown in Table 3.

**Table 3:**
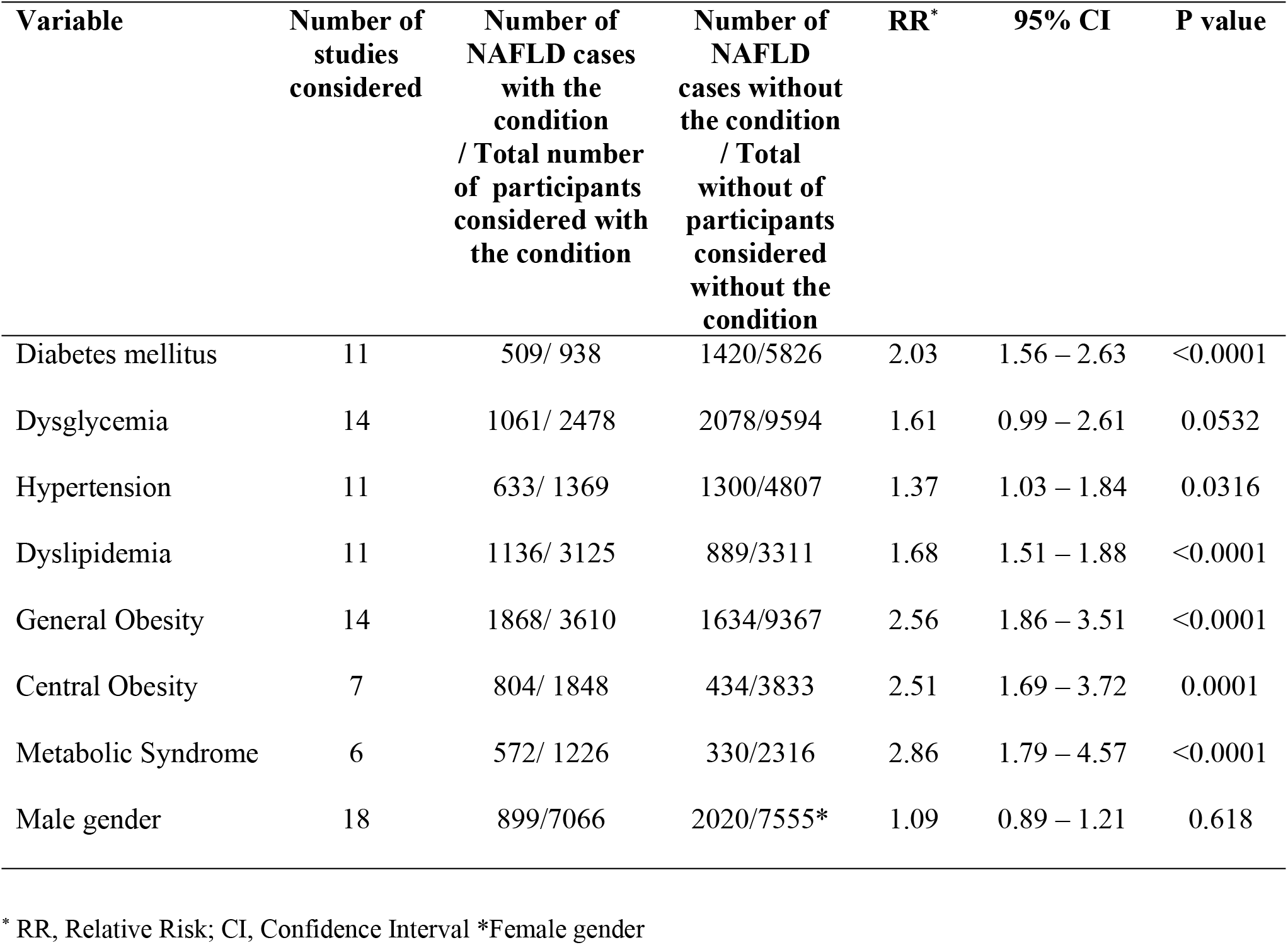
Pooled estimates of relative risk of associated factors with NAFLD in South Asia.

| **Variable** | **Number of studies considered** | **Number of NAFLD cases with the condition / Total number of participants considered with the condition** | **Number of NAFLD cases without the condition / Total without of participants considered without the condition** | **RR\*** | **95% CI** | **P value** |
| --- | --- | --- | --- | --- | --- | --- |
| Diabetes mellitus | 11 | 509/ 938 | 1420/5826 | 2.03 | 1.56 – 2.63 | <0.0001 |
| Dysglycemia | 14 | 1061/ 2478 | 2078/9594 | 1.61 | 0.99 – 2.61 | 0.0532 |
| Hypertension | 11 | 633/ 1369 | 1300/4807 | 1.37 | 1.03 – 1.84 | 0.0316 |
| Dyslipidemia | 11 | 1136/ 3125 | 889/3311 | 1.68 | 1.51 – 1.88 | <0.0001 |
| General Obesity | 14 | 1868/ 3610 | 1634/9367 | 2.56 | 1.86 – 3.51 | <0.0001 |
| Central Obesity | 7 | 804/ 1848 | 434/3833 | 2.51 | 1.69 – 3.72 | 0.0001 |
| Metabolic Syndrome | 6 | 572/ 1226 | 330/2316 | 2.86 | 1.79 – 4.57 | <0.0001 |
| Male gender | 18 | 899/7066 | 2020/7555* | 1.09 | 0.89 – 1.21 | 0.618 |

\* RR, Relative Risk; CI, Confidence Interval \*Female gender

## Discussion

This systematic review and meta-analysis involved 32 and 21 studies, respectively, with 15,455 participants and 4,741 NAFLD cases from five South Asian countries (Bangladesh, India, Nepal, Pakistan and Sri Lanka). We estimated that the pooled prevalence of NAFLD among adults in the general population was 25.2%. Our estimated pooled prevalence of NAFLD in South Asia is comparable to the estimated global prevalence of 25.4%. [1]

Studies in concern showed high heterogeneity where prevalence rates ranged from 8.7% to 73.6%. The overall NAFLD prevalence was 40.4% (95%CI: 33.1% – 47.9%) when all the studies were considered together. Therefore, outlier and influential case diagnostics were performed, and identified the outliers and influence studies. These studies were removed and a meta-analysis was conducted.

Estimating the true prevalence of NAFLD in the general population (with average risk) is a challenge as the disease is mainly asymptomatic. However, this study determined that the overall prevalence of NAFLD among the general population, excluding those with metabolic diseases, was 25.2%. This finding highlights the high proportion of apparently metabolically normal people in the community with NAFLD. There was no difference between prevalence estimates for NAFLD in the current study between males (26.0%) and females (24.7%). Furthermore, residents of rural communities tended to have a similar risk of having NAFLD to residents of urban communities in the South Asian region (26.0% vs 26.6 %, respectively). This finding may reflect a high genetic susceptibility to NAFLD compared to environmental factors for NAFLD in the South Asian region. Therefore, the comparison of NAFLD among residents of urban vs rural communities was essential to look at the influence of environmental factors associated with NAFLD.

In the current meta-analysis, the pooled prevalence of NAFLD among individuals with metabolic diseases (with high risk) was 55.1%, significantly higher than the prevalence in the general population. The pooled prevalence of NAFLD among individuals with metabolic disorders is comparable with the estimated global prevalence of NAFLD among patients with Type 2 diabetes mellitus, 55.5% [15]. The subgroup analysis showed the overall NAFLD prevalence was 46.8 %) in males and 56.3% in females, without significant gender difference in the prevalence rates. Therefore, individuals with metabolic derangements such as general and central obesity, diabetes mellitus, hypertension, and dyslipidaemia are at a considerably higher risk of developing NAFLD than the general population. Furthermore, pooling estimates for the relative risk values of NAFLD showed a significant association with general and central obesity, diabetes mellitus, hypertension, dyslipidaemia, and metabolic syndrome, confirming the strong association between metabolic abnormalities and NAFLD.

A significant proportion of South Asian NAFLD patients are non-obese. This paradox [16, 17] is related to ethnic disparities and visceral fat distribution.[18, 19] In the present study, the pooled estimate for the prevalence of NAFLD in the non-obese population was 11.7%, comparable with prevalence rates reported in western countries.[20, 21] However, the prevalence of non-obese NAFLD among the NAFLD population in the present study was 43.4%, which is far higher than rates reported in western countries.[20, 22, 23] However, the prevalence of non-obese NAFLD in the West and East may not be directly comparable, at least partly due to the different BMI cut-offs, [21] which could have contributed to the dissimilarity observed during the current study. A high index of suspicion is necessary not to miss non-obese NAFLD in clinical practice. It is also noteworthy that non-obese NAFLD is an independent risk factor for coronary artery disease among the South Asian populations. [10, 16, 24]

Before the present study, only a limited number of studies evaluated the prevalence of NAFLD in South Asia. A review article on NAFLD in South Asia in 2016 estimated the epidemiology and determinants of NAFLD in different South Asian countries, including India, Sri Lanka, Bangladesh, Pakistan and Nepal. [9] The prevalence of NAFLD in South Asia was reported to vary from 9% to 45%. The lowest prevalence of NAFLD (8.7-18%) was observed in physically active, poor, lean individuals from rural regions.[25, 26] obesity, acanthosis nigricans, fasting hyperglycaemia, transaminitis, male sex, high BMI, high waist circumference and hypertension were factors that were found to be associated with NAFLD.[26,27] The most recent review article on NAFLD in South Asians, published in 2017, was based on a comprehensive search of available articles on NAFLD in South Asian countries from 1980 to 2016. [8] Estimations of NAFLD across South Asian countries were demonstrated via mapping and tables. A salient finding was that NAFLD was prevalent in cities and rural areas. Among the significant risk factors were age, obesity, insulin resistance and metabolic syndrome. In addition, a higher prevalence of all components of metabolic syndrome was seen among NAFLD cases. The findings reported in these reviews are similar to the present study results.

At the time of the final analysis of our study, a systemic review and meta-analysis of the prevalence of NAFLD in India was published [28]. This study was the first meta-analysis of all published studies on NAFLD in India. The study participants included both children and adults, with an estimated study sample of 26,484 from 50 published studies (children n = 2093, adults n = 23581). While this had a similar methodology to our research, it assessed the NAFLD prevalence within the population sub categorised into average risk groups and into high-risk groups. The high-risk group consisted of obesity, overweight, pre-diabetes, diabetes mellitus, coronary artery disease, metabolic syndrome, obstructive sleep apnoea, polycystic ovarian syndrome and elevated liver enzymes. Notably, the high-risk group had a higher prevalence of NAFLD at 52.8%, comparable to our estimate of 55.1% [28]. Conversely, those in the average-risk group were noted to have a lower prevalence of NAFLD at 28.2% compared to our estimate of 25.2% [28]. Despite the significant heterogeneity among studies and sampling bias, the above findings of the Indian analysis reflect similarly in our study, which found a significant association between NAFLD and metabolic syndrome, general obesity, central obesity, diabetes, dysglycaemia, dyslipidaemia and hypertension. Notably, the high prevalence of NAFLD in India and that of the South Asian region, as noted in our study, further highlights the growing public health concern for the region.

Our study has several strengths and limitations. The present study is the only meta-analysis on the prevalence of NAFLD in the South Asian region. The meta-analysis provides a comprehensive date assessment of the available data over 17 years (2004-2021). We have performed sub-analyses for the general population, rural and urban settings, non-obese populations, and those with metabolic abnormalities. For uniformity, we selected only studies that diagnosed NAFLD based on abdominal ultrasound for the meta-analysis. We carried out an extensive quality assessment of the studies included in the analysis. Only studies with epidemiological studies with satisfactory methodological quality were included to minimise the heterogenicity and risk of bias.

We limited our literature survey to a freely accessible “PubMed” database. We could not include commercially available biomedical literature databases such as Embase due to a lack of funding. A significant limitation of our study is the lack of data from three South Asian countries (Afghanistan, Bhutan and Maldives), limiting our findings’ generalizability beyond the countries included in the analysis. In addition, we could not evaluate age as a risk factor for NAFLD due to most studies’ lack of individual data. Furthermore, this study was limited to available data among adults, excluding the paediatric and adolescent population. According to the reported pooled prevalence of NAFLD in the paediatric and teenage population, [29] it can be predicted that this population would contribute to the rising burden of NAFLD in the South Asian region. However, we could not assess the burden of significant liver fibrosis, which determines the outcomes in patients with NAFLD, as data was not reported in most studies. The high heterogeneity of the included studies was another limitation of the present study. Despite this limitation, the result is likely to represent the NAFLD burden in the South Asian region.

## Conclusion

The prevalence of NAFLD among adults in the South Asian region seems to be comparable to the global average. A considerable proportion are non-obese. NAFLD prevalence was notably higher among individuals with metabolic abnormalities. Diabetes mellitus, hypertension, dyslipidaemia, metabolic syndrome, general and central obesity were identified as associated factors of NAFLD in the South Asian region. Well-designed community-based studies are needed to assess NAFLD prevalence across the South Asian region to plan resource allocation to mitigate the disease burden. Targeted health strategies should be implemented across the South-Asian region to address this growing public health problem.

## Data Availability

All relevant data are within the manuscript and its Supporting Information files.

## Declarations

### Consent statement

Not applicable.

### Ethics Approval

Not applicable

### Availability of data and materials

Data supporting this systematic review and meta-analysis can be only obtained through corresponding author on special request.

### Authors’ contributions

MAN and DSE conceptualised the study. MAN, MYW, SD, DSE and HJdeS formulated the methodology. MAN, MYW, SD and STDeS extracted the data. DSE and SD analysed the data. MAN, SD, DSE drafted the manuscript. STDeS and HJdeS critically analysed and revised the manuscript. All authors read and approved the final manuscript.

## Acknowledgements

Not applicable

## List of abbreviations

BMI: Body Mass Index
CI: Confidence Interval
HDL: High-density lipoproteins
MetD: Metabolic disease
NAFLD: Non-alcoholic fatty liver disease
NASH: Non-alcoholic steatohepatitis
PRISMA: Preferred reporting items for systematic reviews and meta-analysis
RR: Relative Risk
TG: Triglyceride
WC: Waist circumference

